# Validation and Responsiveness of the English version of the Chemotherapy-Induced Alopecia Distress Scale (CADS) in Breast Cancer Patients

**DOI:** 10.1101/2023.11.05.23298093

**Authors:** L. Kraehenbuehl, D. Kang, A. S. Bang, K. F. Ketosugbo, J. Hay, Sujata Patil, S. Goldfarb, J. Cho, M. E. Lacouture

## Abstract

**Purpose:** This study aimed to validate the chemotherapy-induced alopecia distress scale (CADS) in a diverse English-speaking population and patients with endocrine treatment- induced alopecia (EIA).

**Objective:** Chemotherapy and endocrine therapy commonly cause alopecia in breast cancer patients, leading to significant psychological and social challenges. The CADS was developed to assess the psychosocial impact of alopecia, but its generalizability beyond Korean patients requires further investigation.

**Methods:** Data from the CHANCE study (NCT02530177), which focused on non-metastatic breast cancer, was used. The cohort included 256 patients, and CADS data were collected at baseline, six months after chemotherapy completion, or 12 months after initiating endocrine therapy. The CADS questionnaire comprised 17 items covering physical and emotional health, daily activities, and relationships. Reliability was assessed using Cronbach’s alpha, and responsiveness was measured by effect size.

**Results:** The CADS exhibited good reliability, with a Cronbach’s alpha of 0.91 for the overall score, indicating acceptable internal consistency in both chemotherapy (0.89) and endocrine therapy (0.86) groups. Longitudinal responsiveness was supported by an effect size of 0.49 between decreasing satisfaction with hair growth and increasing emotional distress. Cross-sectional validity was confirmed, with effect sizes of 0.91 and 0.92 for satisfaction with hair growth and emotional and activity domains, respectively.

**Conclusion:** The CADS is a valid and responsive tool for assessing the psychosocial impact of chemotherapy-induced alopecia and endocrine treatment-induced alopecia in a diverse Western patient population.

## BACKGROUND

Alopecia induced by cancer treatment can be one of breast cancer patients’ most stressful and burdensome adverse events. The incidence of alopecia in early-stage breast cancer patients undergoing cytotoxic chemotherapy is expected to be above 80% with taxanes and 60-100% with anthracyclines^1^. Persistent chemotherapy-induced-alopecia (pCIA) is defined as incomplete hair regrowth six months after completion of chemotherapy and occurs in 39.5% of breast cancer patients^2^. Additionally, 22.4% and 31.8% of women undergoing adjuvant endocrine therapy for breast cancer report hair loss and hair thinning, respectively^1^. The multi-faceted importance of CIA has recently been reviewed^3^, emphasizing the importance of CIA as a representation of illness to the patient and the public.

Previous studies have shown that alopecia can affect treatment choice and dose intensity and lead to early treatment discontinuation. Alopecia can cause disproportionate psychosocial impairment, affecting patients’ functional status, emotional well-being, and relationships with others. It has been reported as the third most burdensome adverse event in treating gynecological- and breast cancer. ^4–8^. Patients with higher distress related to cancer treatment- associated alopecia are 1.5 times more likely to suffer from depression than those with lower distress^9^.

To our knowledge, the incidence of chemotherapy-associated alopecia in men and potential ethnic or racial differences in women have not been studied in English literature. Further research needs to be conducted to understand better the significance of ethnicity, gender, and race in patients experiencing alopecia from cancer therapy. In most clinical trials, as well as in routine oncological practice, grading of alopecia is based on the Common Terminology Criteria for Adverse Events (CTCAE), distinguishing between no hair loss (grade 0), “hair loss of <50% of normal for that individual that is not obvious from a distance but only on close inspection; a different hairstyle may be required to cover the hair loss, but it does not require a wig or hair piece to camouflage” (grade 1) and “hair loss of >=50% normal for that individual that is readily apparent to others; a wig or hairpiece is necessary if the patient desires to camouflage the hair loss completely; associated with psychosocial impact” (grade 2)^10^. In addition, a patient-reported outcome grading tool (PRO-CTCAE) is available as a companion measurement. Patients are requested to quantify their hair loss during the last seven days as ‘not at all’ (grade 0), ‘a little bit’ (grade 1), ‘somewhat’ (grade 2), ‘quite a bit’ (grade 3) and ‘very much’ (grade 4)^11^. The CTCAE grading is well-recognized and easy to use but does not capture the psychological impact of alopecia in cancer patients.

Some tools assess the impact of hair disorders on quality of life, including Hairdex^12^, the modified Dermatology Life Quality Index (DLQI), adapted DLQI^13^, and additional tools ^14^ have been proposed. However, no tools specifically cater to patients experiencing CIA or EIA outside the Chemotherapy-induced Alopecia Distress Scale (CADS). Recently, the CADS^15^, measuring CIA-induced distress in patients with breast cancer, was developed and introduced by Cho and colleagues^16^. CADS is the first validated tool to quantify the psychological impact of CIA on breast cancer patients. Testing 25 items, 17 items, covering the four domains ‘physical’ (2), ‘emotional’ (6), ‘activity’ (6), and ‘relationship’ (3) were identified through exploratory factor analysis.

CADS was validated in a cohort of 305 Korean breast cancer patients and found reliable and valid, with a coefficient alpha of 0.95 overall and 0.77-0.95 for the subdomains. Total scores ranging from 0 to 51 and mean scores by domains were recorded. Higher scores represent higher levels of distress. The availability of a total score allows for a simple understanding of a patient’s alopecia-related distress. Subsequently, the values from each domain provide a more detailed assessment of the experienced distress. It has been recognized as a crucial tool in monitoring the CIA’s impact on cancer patients. It has been translated and validated for Chinese patients^17^ but not for English-speaking populations. Even more importantly, CADS has not been validated for EIA, and a comparable tool for this population is missing. The initial validation study’s design could not assess responsiveness^18^, which is crucial in understanding the longitudinal impact on patients over time^19^ and constitutes an essential aspect of validitation^20^. We used a prospective cohort of patients receiving chemotherapy or endocrine therapy for non-metastatic breast cancer^21^ to validate the English version of the CADS scale. ^21^ In addition, we aim to assess the responsiveness of the CADS, which was not evaluated in the original study due to study design limitations^19^.

## METHODS

### Participants

We used cohort data from a single-center, prospective cohort study entitled *Chemotherapy- Induced Hair Changes and Alopecia, Skin Aging and Nail Changes in Women with Non- Metastatic Breast Cancer* (CHANCE study NCT02530177^21^). The CHANCE study was conducted at Memorial Sloan Kettering Cancer Center to quantify the incidence of persistent alopecia and alopecia in breast cancer patients receiving cytotoxic chemotherapy and endocrine therapy, respectively. Subjects were eligible to participate if they were diagnosed with breast cancer (ductal carcinoma in situ (DCIS), stage I to III), had no sign of metastasis, and were expected to have cytotoxic chemotherapy and/or endocrine therapy. The study was designed to enroll 100 patients in each of the five treatment cohorts for 500 patients who were followed over time. Due to the nature of the various treatment types and individual susceptibility, this included patients with G0-G2 alopecia.

Since the purpose of this study was the validation of CADS in patients with chemotherapy and/or endocrine therapy, we used 256 patients with chemotherapy and/or endocrine therapy who have completed their study visit at the completion of chemotherapy in patients with chemotherapy, and at week 24 from baseline in patients with endocrine treatment only. To test reliability, we used visits at 24 months and 30 months after enrollment expecting only a slight change in the chemotherapy-induced alopecia distress and patients who did not change hair status between the period. This number is lower than the target enrolment as patients from the control cohort (pre-and post-menopausal women, assessed at baseline only) were not evaluated, and patients with missing data points at either baseline or specified endpoint were excluded.

All procedures were approved by the Institutional Review Board of Memorial Sloan Kettering Cancer Center, New York, NY (IRB Number: 15-198 A(10), NCT02530177), and all participants provided written informed consent before participation.

### Measurement

The CADS is a psychometric scale for assessing the distress that breast cancer patients experience because of CIA. Its reliability and validity were established with a cross- sectional survey of 305 Korean women with breast cancer.^16^ Exploratory factor analysis and confirmatory factor analysis yielded 17 items in four domains with good model fit, including physical (two questions), emotional (six questions), daily activity (six questions), and relationship (three questions) domains. The response is based on a 4-point Likert scale (0 = not at all, 1 = a little, 2 = quite a bit, and 3 = very much). The CADS total score is calculated by summing responses for all items, ranging from 0 to 51. A higher score indicates higher distress. The English version of CADS used in this study has been translated and linguistically validated by Dr. Cho (JC) using the standardized methodology recommended by Functional Assessment of Chronic Illness Therapy multilingual translation (FACITtrans) ^22^. JC’s research team reviewed all documentation pertaining to the CADS translation, including the item history and decisions about item rephrasing, before being finalized for cognitive testing. No translation issues related to the response options, linguistics, and conceptual equivalence to the original CADS measure, as intended, were reported. The participants subsequently underwent cognitive debriefing for 30 min to evaluate comprehension; ease of response; and acceptability of the terminology, phrasing, and response options. Cognitive interviews were conducted by one oncology nurse and a behavioral scientist. Cancer patients were recruited until saturation; 5 participated in the cognitive interviews. The cognitive debriefing revealed that the participants generally comprehended the CADS well.

We also evaluated the association between the CADS and standardized phototrichogram data to confirm the construct validity (“known groups”). The objective data included the number and thickness of hair and patients’ reported satisfaction with hair. Clinical characteristics such as demographic information of study participants were obtained from electronic medical records.

### Statistical analysis

#### Item internal consistency

Descriptive statistics were used to report participants’ characteristics and the mean and standard deviation (SD) of each item of the CADS. We calculated Cronbach’s alpha to test the internal consistency and reliability of the CADS. We expected a value greater than 0.75, which is the standard for defining the acceptable reliability of an instrument. At least 209 patients were required to estimate Cronbach’s alpha of 0.75 with ±5% accuracy at 95% confidence intervals in the 17-item CADS. ^23^ This study had 256 patients, which was sufficient. To further evaluate the test’s internal consistency, we also estimated the corrected item-rest correlation, which represents the correlation of the item with the rest of the total score. Items exceeding a correlation of 0.40 are judged as ‘good.’^24^

We initially attempted to use structural equation modeling (SEM) for the CFA but encountered difficulties constraining secondary loadings to zero without compromising model fit. In addition, because treating ordinal data as continuous in SEM would not be appropriate, we adopted exploratory structural equation modeling (ESEM), which allows for an unconstrained measurement model with all possible cross-loadings as free parameters. ^25^ This approach is identified through the specification of a rotation option. It provides standard errors and fit statistics for freely estimated parameters, making it a suitable method for our analysis in this study.

Several goodness-of-fit indices were used to evaluate the model fit, including the comparative-fit-index (CFI), Tucker-Lewis Index (TLI), standardized root-mean-squared residual (SRMR), and root mean square error of approximation (RMSEA). A CFI >0.9, TLI >0.9, SRMR <0.08, and RMSEA <0.06 indicate a good fit for the data.^26^

#### Known-Group Validity

We determined cross-sectional and longitudinal known-group validity using variables of patients with and without chemotherapy and objective measures including the number of hair and thickness of hair. Cross-sectional known-group validity independent t-tests were used to evaluate whether CADS scores were different based on chemotherapy. In addition, the correlation between CADS with the number of hair and thickness of hair shafts were assessed using Pearson’s correlation, and we expected total scores to at least moderately correlate (0.30≤***r***≥0.70).

#### Reliability

The test-retest reliability of the CADS was measured using the intra-class correlation coefficient (ICC) using a two-way mixed model. A questionnaire is considered reliable at ICC values> 0.70.^26^

#### Responsiveness

Responsiveness is the capacity of the questionnaire to identify possible changes in the construct associated with the clinical condition over time.^28^ This responsiveness was measured by the effect size (ES).^29^ ES was calculated by the variation of the score in the CADS between patients with and without satisfaction with hair growing or decreasing and no change of satisfaction with hair growing by the standard deviation of the score. Based on the previous studies^9,30^, the following specific hypotheses were tested: 1) decreasing satisfaction with hair growth would have a positive correlation of at least 0.5 with the increasing emotional domain in CADS after treatment; 2) decreasing satisfaction with hair growth would have a positive correlation of at least 0.2 with the increasing physical, activity, and relationship domains in CADS after treatment; and 3) decreasing satisfaction with hair growth will have a positive correlation of at least 0.2 with the increasing total score of CADS after treatment.

The significance level was at *P<0.05* (two-sided), and all statistical analyses were done using STATA software package 16 (STATA Corp., 4905 Lakeway Drive, College Station, TX 77845 USA).

## RESULTS

### Study participants

The mean age was 53.1 years, and 30% were BC stage II or III. Among the study participants, 57.4% received chemotherapy (Table 1). On a scale of 0 to 51, the mean CADS score was 3.1 (SD = 5.3) and 5.9 (SD = 7.1) at baseline and cycle 8, respectively.

**Table 1.** Patient characteristics and baseline measurements (N= 256)

| Characteristics | Overall<br>(N= 256)<br>N (%) | Sub-cohort |  |
| --- | --- | --- | --- |
|  |  | Chemotherapy<br>(N = 147) | Endocrine<br>therapy<br>only<br>(N = 109) |
| **Age** |  |  |  |
| Mean (SD), | 53.1 (11.4) | 50.9 (11.5) | 56.2 (10.6) |
| **Race (N = 216)** |  |  |  |
| White | 151 (69.9) | 81 (66.4) | 70 (74.5) |
| Black or African American | 21 (9.7) | 15 (12.3) | 6 (6.4) |
| Latino/ Hispanic | 16 (7.4) | 12 (9.8) | 4 (4.3) |
| Asian | 17 (7.9) | 9 (7.4) | 8 (8.5) |
| Others | 11 (5.1) | 5 (4.1) | 6 (6.4) |
| **Disease stage at diagnosis** |  |  |  |
| DCIS | 18 (7.0) | 0 | 18 (16.5) |
| I | 157 (61.3) | 77 (52.4) | 80 (73.4) |
| II | 71 (27.7) | 60 (40.8) | 11 (10.1) |
| III | 9 (3.5) | 9 (6.1) | 0 |
| Unknown | 1 (0.4) | 1 (0.7) | 0 |
| **Treatment modality** |  |  |  |
| **Cytotoxic chemotherapy** |  |  |  |
| ddAC-T | 73 (28.5) | 73 (49.7) |  |
| CMF | 48 (18.8) | 48 (32.7) |  |
| Newer Combination Regimens | 26 (10.2) | 26 (17.7) |  |
| **Endocrine therapy** |  |  |  |
| Tamoxifen | 46 (18.0) |  | 46 (42.2) |
| Aromatase Inhibitor | 63 (24.6) |  | 63 (57.8) |

\*Patient who received chemotherapy

### Item internal consistency

The Cronbach’s alpha coefficients of 4 sub-domains in the CADS ranged from 0.50 to 0.88, indicating satisfactory internal consistency except for physical function. When any items were removed, item-rest correlations varied from 0.18 to 0.74. While all the items had generally accepted levels of item-rest correlation (≥0.40), the item “The area is itching” (r = 0.18), “The area is burning or prickling resulting pain.” (r = 0.31), and “I have difficulty doing personal care such as bath and make-up.” (r = 0.35) had a relatively low correlation with other items in the domain (Table 2). However overall CADS scale of Cronbach’s alpha coefficients was 0.91, indicating good reliability. The Cronbach’s alpha coefficients were 0.89 and 0.86 in patients on chemotherapy and patients on endocrine therapy only, respectively, and the CADS had acceptable reliability in both groups.

**Table 2.** Reliability of the Chemotherapy-Induced Alopecia Distress Scale (N= 256) Chemotherapy induce Alopecia Scale, CADS

### Confirmatory factor analysis

In confirmatory factor analysis, test statistics, degrees of freedom, and p values of the Chi- square test were 2384.075, 136, and <0.01. The goodness-of-fit indices for CADS (Figure 1) were relatively high (CFI = 0.940, TLI = 0.890, SRMR = 0.041, and RMSEA = 0.084 (95% CI = 0.071, 0.098)).

**Figure 1.**
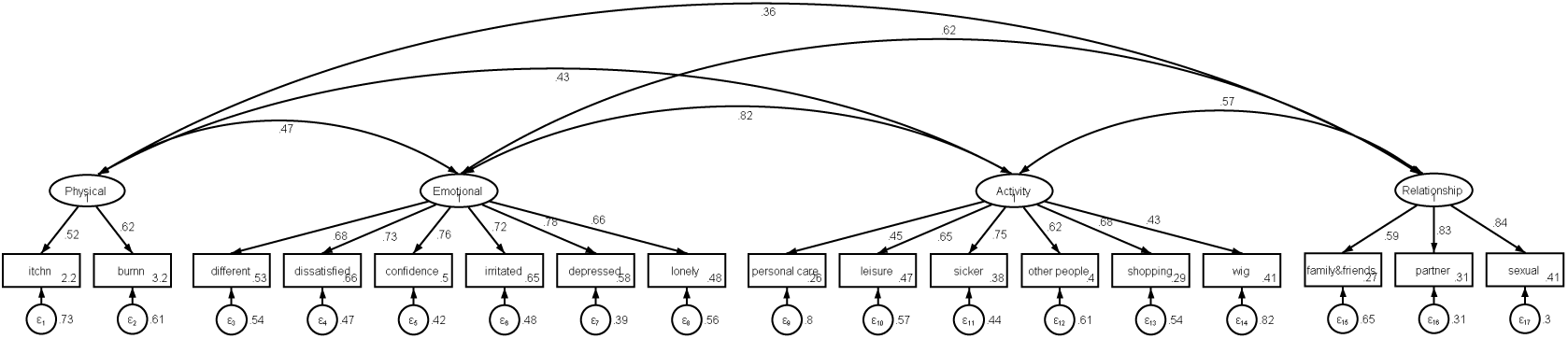
Confirmatory factor analysis of Chemotherapy-induced Alopecia Distress Scale items.

### Cross-sectional Known-Group Validity

Compared to patients only on endocrine therapy, patients on chemotherapy were more likely to have higher distress due to CIA (Table 2), and the difference was statistically significant (P < 0.05). Overall, the highest distress item was “I am dissatisfied with my appearance.^”^ followed by “I always wear a wig or scarf to hide hair loss” (Table 2). The highest distress items in the chemotherapy- and endocrine therapy group were “I always wear a wig or scarf to hide hair loss” and “I am easily irritated and stressed,” respectively (Table 2).

In cross-sectional known group analysis using subjective measurement, the ES between the with and without satisfaction with hair growth and higher emotional and activity domain was 0.91 and 0.92, respectively. The satisfaction was strongly associated with a total score of CADS (ES = 0.97) (Table 3). The differences in CADS score between patients with and without satisfaction with hair growth were observed to be of large ES in both chemotherapies (ES = 0.60) and endocrine therapy-only group (ES = 0.52) (Table 3).

**Table 3.**
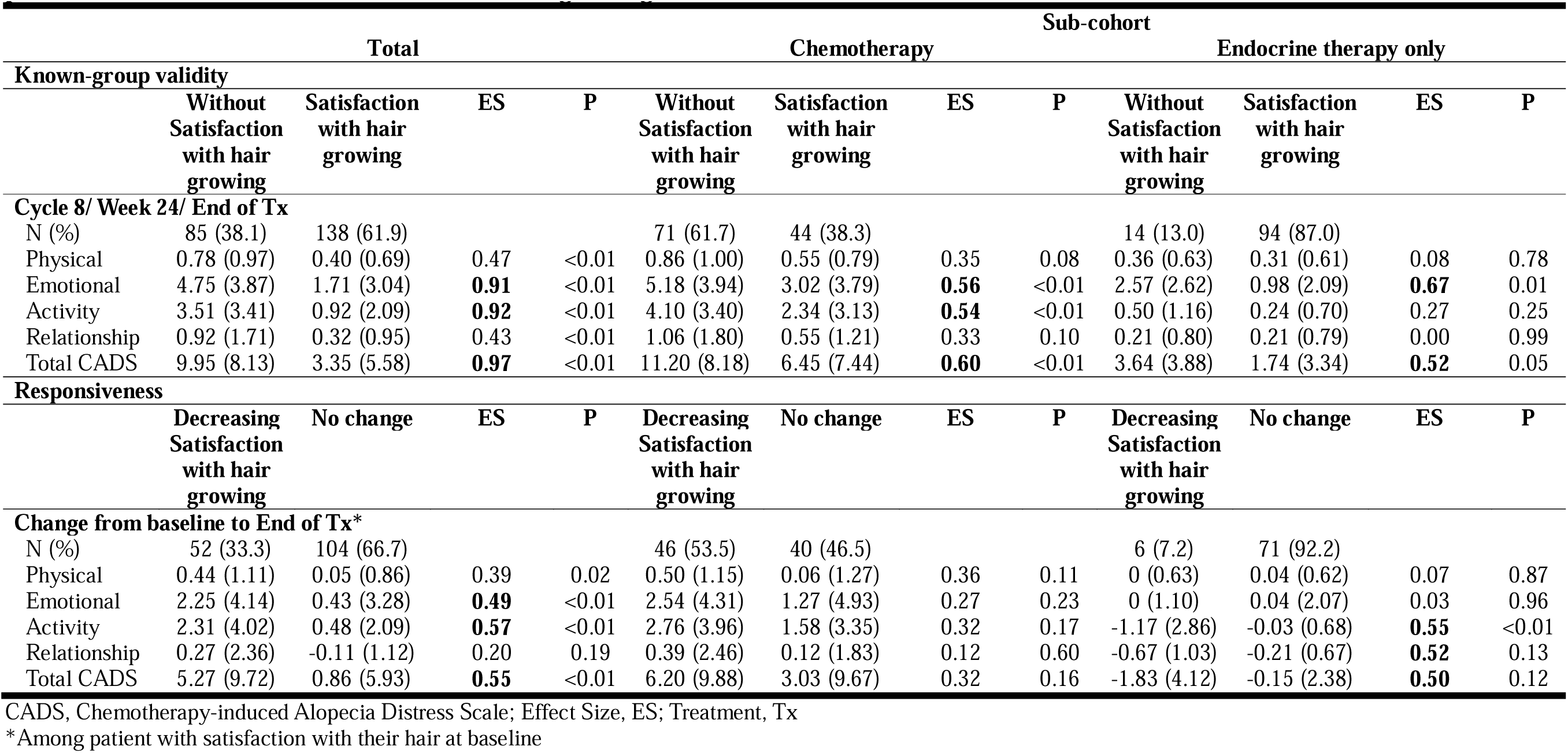
Known-group validity and Responsiveness validity of Chemotherapy-induced alopecia distress scale (CADS) subscale by patients with and without satisfaction with hair growing baseline and after treatment.

Regarding the objective phototrichogram data, a moderate correlation was observed between the total score of CADS, number of hair (r = −0.33), and thickness of hair shafts (r = −0.44) at the completion of chemotherapy or 24 weeks after initiation of endocrine therapy. Among the domains, the activity domain was more likely to have a higher correlation with the number of hair (r = −0.40) and thickness of hair shafts (r = −0.49) (Table 4). The correlation between CADS score and the number and thickness of hair was observed to be of large ES in both chemotherapies (ES = 0.60) and endocrine therapy-only group (ES = 0.52) (Table 3).

**Table 4.** Cross-sectional and Longitudinal correlation coefficients between scores of the Chemotherapy-Induced Alopecia Distress Scale (CADS), and number and thickness of hair before and after chemotherapy.

|  | Total (N= 257) |  | Sub-cohort |  |  |  |
| --- | --- | --- | --- | --- | --- | --- |
|  | Number of hair<br>(N/cm<sup>2</sup>) | Thickness of hair<br>(mm) | Chemotherapy |  | Endocrine therapy only |  |
|  |  |  | Number of hair<br>(N/cm<sup>2</sup>) | Thickness of hair<br>(mm) | Number of hair<br>(N/cm<sup>2</sup>) | Thickness of hair<br>(mm) |
| **Cross-sectional score** |  |  |  |  |  |  |
| Physical | -0.15** | -0.12* | -0.02 | -0.03 | -0.17 | -0.06 |
| Emotional | -0.27** | **-0.36\*\*** | -0.05 | -0.23** | -0.14 | -0.00 |
| Activity | **-0.40\*\*** | **-0.49\*\*** | -0.18* | **-0.34\*\*** | -0.14 | 0.02 |
| Relationship | -0.11 | -0.23** | -0.00 | -0.18* | 0.02 | 0.04 |
| Total CADS | **-0.33\*\*** | **-0.44\*\*** | -0.10 | **-0.30\*\*** | -0.15 | 0.02 |
| **Longitudinal change of score** |  |  |  |  |  |  |
| Physical | -0.06 | -0.03 | 0.00 | 0.05 | 0.07 | 0.04 |
| Emotional | -0.20** | -0.29** | -0.08 | -0.22* | -0.06 | 0.09 |
| Activity | -0.27** | **-0.39\*\*** | -0.11 | -0.28** | 0.11 | 0.24* |
| Relationship | -0.07 | -0.26** | -0.01 | -0.24* | 0.29** | 0.26 |
| Total CADS | -0.23** | **-0.35\*\*** | -0.08 | -0.26** | 0.11 | 0.20 |

\* P < 0.05
\*\*P < 0.01

### Longitudinal Reliability and Responsiveness

Among the patients who did not change hair status between 24 months and 30 months after enrollment, the ICC of CADS total score between 24 months and 30 months was 0.83 (95% CI 0.73, 0.89), which indicated satisfactory consistency.

In responsiveness based on the satisfaction with hair growing, the results in relation to our hypotheses were as follows: 1) The ES between the decreasing satisfaction with hair growing and increasing emotional domain was 0.49, which was in accordance with hypothesis 1. 2), 3), and 4) The ES between the decreasing satisfaction and the average change score of the physical, activity and relationship was 0.39, 0.57 and 0.20, which was in accordance with hypotheses 2,3, and 4. 5) The correlation between decreasing satisfaction with hair growing and CADS total score was 0.55, which was in accordance with hypotheses 5) (Table 3). There was moderate ES between decreasing satisfaction and change of CADS score in both chemotherapy (ES = 0.30) and endocrine therapy-only group (ES = 0.50) (Table 3).

The correlation between the change in the total score of CADS and the difference in the thickness of hair (r = −0.35) the observed correlation was moderate. The change in the activity domain was also observed to have the highest correlation with the change in objective measures (Table 4). Between the shift in the number and thickness of hair and the change in CADS score, a moderate correlation was observed in chemotherapy (ES = 0.26), but a weak correlation in the endocrine therapy-only group (Table 4).

## Discussion

To fill an essential gap in measuring psychological distress associated with alopecia in cancer patients, we validated the previously established CADS in a diverse English-speaking population at Memorial Sloan Kettering Cancer Center in the United States. Additionally, we aimed to expand the use of CADS to include patients treated with endocrine therapies. The CADS, as demonstrated in this study, is reliable for administration in this non-Korean patient population and can be used in patients with EIA in addition to CIA patients (Cronbach’s alpha 0.86 and 0.89). Furthermore, we identified a moderate inverse correlation between hair count and thickness assessed by phototrichogram with CADS. This further supports the validity of CADS.

Interestingly, this study’s mean total CADS score at baseline and completion of chemotherapy or 24 weeks after initiation of endocrine therapy was 3.1 and 5.9, respectively. These total scores are considerably lower than the median of 14 reported in the initial CADS validation study^18^. Possible explanations for this finding include the inclusion of patients with EIA, which tends to be less severe and leads to less psychological distress, and cultural differences in the perception of female hair loss in general and during cancer treatment.

In terms of validity and reliability, all items had moderate or high loadings (>0.5), supporting the factorability of the items in a development study. However, in our study, two items in the physical domain revealed low loadings (*“The area is itching”* and *“The area is burning or prickling, resulting in pain”*). This may be due to the status of our study participants. Patients typically describe hot, itchy, tender, or tingly sensations on their scalp (trichodynia) when actively losing hair from chemotherapy. However, in this study, CADS was assessed at completion of chemotherapy when hair was expected to regrow. This may explain the lower values in these items. To ensure the consistency of the CADS as an international standardized instrument for psychometric analysis, we decided to maintain the aforementioned items. Further studies will be required to modify these items and their responsiveness for patients after completion of chemotherapy.

Regarding convergent validities using known group analysis, we found that dissatisfaction with hair growing was associated with a higher total score of CADS (ES = 0.97). Emotional and activity stress domains were higher in patients who had dissatisfaction with hair growth. In addition, we also found that with the objective phototrichogram data, a moderate correlation was observed between the total score of CADS, number of hair (r = - 0.33), and thickness of hair shafts at completion of chemotherapy or 24 weeks after initiation of endocrine therapy. The US Food and Drug Administration (FDA) has noted the need to develop endpoints for clinical trials to measure aspects of alopecia that are important to patients.^27^ Considering the CADS was associated with hair density and hair thinning, which was objectively quantified, as well as patient satisfaction, CADS could be a helpful measurement tool as a valid patient-reported outcome response scale in clinical trials.

In responsiveness, the correlation between decreasing satisfaction with hair growing and CADS total score was 0.55, and the minimal clinically meaningful difference of CADS could be 5 points at complete chemotherapy. This means that in individual patients, a change of 5 points, considered important by patients, cannot be distinguished from measurement error with 95% confidence, so changes in individual patients should be interpreted with caution.

The initial development of CADS has been a significant advancement in quantifying the psychological impact of CIA^16^. However, using a questionnaire in different cultural contexts and patient settings must be validated to ensure that the questionnaire applies to the patients^28^. This also implies that caution should be warranted when using CADS in similar but not identical populations, e.g., western European patients in the future. An additional potential limitation is the minor logistical burden associated with the administration and scoring of the CADS, as well as time constraints for patients when used outside of clinical trials. However, the ability to quantify the psychological distress of patients experiencing chemotherapy-associated alopecia longitudinally will allow for better-shared decision-making regarding alopecia treatment, prevention, and psychological support. The questionnaire’s advantages benefit patients overall and outweigh the logistical burdens.

### Limitations of this study

Limitations include single-center design. Further studies should validate CADS in different patient populations with various cultures and languages and consider conducting international multi-center studies. Second, we did not conduct a test-retest. However, the development study^15^ and other validation study^17^ have already proven the reliability of the CADS, and we expected our results to be similar to those in previous studies. Furthermore, when we performed a reliability test using patients who did not experience a change in their hair condition between two visits when the chemotherapy-induced alopecia status was relativity consistent, the ICC was satisfied with the cut-off (ICC = 0.83). Lastly, since the item was a categorical variable, the statistical interpretation was limited because approximate fit indexes were initially developed for continuous outcomes. ^29^ However, studies support that ordinal variables could be considered continuous variables in the structural equation model.^29^ and the model’s goodness of fit in this study are acceptable.

### Clinical implications

The validated CADS is the first tool established for a diverse patient population in the United States. Furthermore, including EIA patients expands this tool’s validation to another patient population.

### Conclusions

We have validated CADS as a reliable and responsive measurement of psychological distress due to both CIA and EIA in a diverse Western population. We encourage its wide use in research and clinical practice. Validation of CADS in additional linguistic, cultural, and ethnic contexts will allow more patients to benefit from this psychometric tool.

## Data Availability

All data produced in the present study are available upon reasonable request to the authors

## Acknowledgments

Funding was provided by the RJR fund

NIH/NCI Cancer Center Support Grant P30 CA008748

Lukas Kraehenbuehl was also supported by the Swiss National Science Foundation P400PM_199318.

## REFERENCE

1 Trüeb RM. Chemotherapy-induced alopecia. Seminars in cutaneous medicine and surgery. 28. No longer published by Elsevier; 2009:11–14.

2 Kang D, Kim IR, Choi EK et al. Permanent Chemotherapy-Induced Alopecia in Patients with Breast Cancer: A 3-Year Prospective Cohort Study. Oncologist 2019; 24 (3): 414–420.

3 Boland V, Brady A-M, Drury A. The physical, psychological and social experiences of alopecia among women receiving chemotherapy: An integrative literature review. European Journal of Oncology Nursing 2020; 49: 101840.

4 McGarvey EL, Baum LD, Pinkerton RC et al. Psychological Sequelae and Alopecia Among Women with Cancer. Cancer Practice 2001; 9 (6): 283–289.

5 Freites-Martinez A, Shapiro J, Chan D et al. Endocrine Therapy–Induced Alopecia in Patients With Breast Cancer. JAMA Dermatology 2018; 154 (6): 670–675.

6 Moscetti L, Agnese Fabbri M, Sperduti I et al. Adjuvant aromatase inhibitor therapy in early breast cancer: what factors lead patients to discontinue treatment? Tumori 2015; 101 (5): 469–473.

7 Gallicchio L, Calhoun C, Helzlsouer KJ. Aromatase inhibitor therapy and hair loss among breast cancer survivors. Breast Cancer Res Treat 2013; 142 (2): 435–443.

8 Lemieux J, Maunsell E, Provencher L. Chemotherapy-induced alopecia and effects on quality of life among women with breast cancer: a literature review. Psycho-oncology 2008; 17 (4): 317–328.

9 Choi EK, Kim IR, Chang O et al. Impact of chemotherapy-induced alopecia distress on body image, psychosocial well-being, and depression in breast cancer patients. Psycho-oncology 2014; 23 (10): 1103–1110.

10 CTEP N. CTCAE v5.0. 2020. https://ctep.cancer.gov/protocoldevelopment/electronic_applications/docs/CTCAE_v5_Quick_Reference_8.5x11.pdf.

11 Basch E, Reeve BB, Mitchell SA et al. Development of the National Cancer Institute’s Patient-Reported Outcomes Version of the Common Terminology Criteria for Adverse Events (PRO-CTCAE). JNCI: Journal of the National Cancer Institute 2014; 106 (9).

12 Fischer TW, Schmidt S, Strauss B et al. HairdexEin Instrument zur Untersuchung der krankheitsbezogenen Lebensqualität bei Patienten mit Haarerkrankungen. Der Hautarzt 2001; 52 (3): 219–227.

13 Williamson D, Gonzalez M, Finlay AY. The effect of hair loss on quality of life. J Eur Acad Dermatol Venereol 2001; 15 (2): 137–139.

14 Davis DS, Callender VD. Review of quality of life studies in women with alopecia. International Journal of Women’s Dermatology 2018; 4 (1): 18–22.

15 Cowley L, Heyman B, Stanton M et al. How women receiving adjuvant chemotherapy for breast cancer cope with their treatment: a risk management perspective. J Adv Nurs 2000; 31 (2): 314–321.

16 Cho J, Choi EK, Kim IR et al. Development and validation of Chemotherapy-induced Alopecia Distress Scale (CADS) for breast cancer patients. Ann Oncol 2014; 25 (2): 346–351.

17 Cong W, Wu Y, Liu L et al. A Chinese version of the chemotherapy-induced alopecia distress scale based on reliability and validity assessment in breast cancer patients. Support Care Cancer 2020; 28 (9): 4327–4336.

18 Cho J, Choi EK, Kim IR et al. Development and validation of Chemotherapy-induced Alopecia Distress Scale (CADS) for breast cancer patients. Annals of Oncology 2014; 25 (2): 346–351.

19 Mokkink LB, Terwee CB, Patrick DL et al. The COSMIN study reached international consensus on taxonomy, terminology, and definitions of measurement properties for health-related patient-reported outcomes. J Clin Epidemiol 2010; 63 (7): 737–745.

20 de Boer MR, Terwee CB, de Vet HCW et al. Evaluation of Cross-sectional and Longitudinal Construct Validity of Two Vision-related Quality of Life Questionnaires: The LVQOL and VCM1. Quality of Life Research 2006; 15 (2): 233–248.

21 Lacouture ME, Freites-Martinez A, Patil S et al. The CHANCE study: A prospective, longitudinal study of chemotherapy and hormonal therapy induced hair changes and alopecia, skin aging and nail changes in women with non-metastatic breast cancer. Journal of Clinical Oncology 2018; 36 (15_suppl): e12500-e12500.

22 Lent L, Hahn E, Eremenco S et al. Using cross-cultural input to adapt the Functional Assessment of Chronic Illness Therapy (FACIT) scales. Acta oncologica (Stockholm, Sweden) 1999; 38 (6): 695–702.

23 Bonett DG. Sample Size Requirements for Testing and Estimating Coefficient Alpha. Journal of Educational and Behavioral Statistics 2002; 27 (4): 335–340.

24 Aiken LR. Book Reviews : Measuring Educational Achievement by Robert L. Ebel. Englewood Cliffs, N. J.: Prentice-Hall, 1965. pp. xii + 481. Educational and Psychological Measurement 1965; 25 (4): 1167–1169.

25 van Zyl LE, Ten Klooster PM. Exploratory Structural Equation Modeling: Practical Guidelines and Tutorial With a Convenient Online Tool for Mplus. Front Psychiatry 2021; 12: 795672.

26 de Souza JA, Yap BJ, Wroblewski K et al. Measuring financial toxicity as a clinically relevant patient-reported outcome: The validation of the COmprehensive Score for financial Toxicity (COST). Cancer 2017; 123 (3): 476–484.

27 Food U, Administration D. The voice of the patient. A series of reports from the US Food and Drug Administration’s (FDA’s) Patient-Focused Drug Development Initiative. Lung Cancer Available from: http://waybackarchive-itorg/7993/20171114193841/https://www.fdagov/ForIndustry/UserFees/PrescriptionDrugUserFee/ucm353273htm Accessed February 2018; 28.

28 Tsang S, Royse CF, Terkawi AS. Guidelines for developing, translating, and validating a questionnaire in perioperative and pain medicine. Saudi J Anaesth 2017; 11 (Suppl 1): S80–s89.

29 Muthén B. A general structural equation model with dichotomous, ordered categorical, and continuous latent variable indicators. Psychometrika 1984; 49 (1): 115–132.

